# Cortical complexity analyses and their cognitive correlate in Alzheimer’s disease and frontotemporal dementia

**DOI:** 10.1101/19013466

**Authors:** Nicolas Nicastro, Maura Malpetti, Thomas E. Cope, William Richard Bevan-Jones, Elijah Mak, Luca Passamonti, James B. Rowe, John T. O’brien

**Author notes:** **CORRESPONDENCE:** Dr N. NICASTRO, MD, Division of Neurology, Department of Clinical Neurosciences, Geneva University Hospitals, 4, rue G. Perret-Gentil, 1205 Geneva, SWITZERLAND. co-senior authors (James B. Rowe, John T. O’brien).

## Abstract

The changes of cortical structure in Alzheimer’s disease (AD) and frontotemporal dementia (FTD) are usually described in terms of atrophy. However, neurodegenerative diseases may also affect the complexity of cortical shape, such as the fractal dimension of the brain surface. Thirty-two people with symptomatic AD-pathology (clinically probable AD, n=18, and amyloid-positive mild cognitive impairment, n=14), 24 with FTD and 28 healthy controls underwent high-resolution 3T structural brain MRI. Using surface-based morphometry, we created vertex-wise cortical thickness and fractal dimension maps for group comparisons and correlations with cognitive measures in AD and FTD. In addition to the well-established pattern of cortical thinning encompassing temporoparietal regions in AD and frontotemporal areas in FTD (FDR p< 0.05), we observed reductions of fractal dimension specifically involving precuneus and posterior cingulate for AD and orbitofrontal cortex and insula for FTD. Correlational analyses between fractal dimension and cognition showed that these regions were particularly vulnerable with regards to memory and language impairment in both AD and FTD. This study demonstrates a distinct pattern of fractal dimension impairment and correlation with cognition. Further studies are required to assess cortical complexity measures at earlier disease stages (e.g. in prodromal/ asymptomatic carriers of FTD-related gene mutations) to assess whether fractal dimension represents a sensitive imaging marker for prevention and diagnostic strategies.

## 1. INTRODUCTION

Considering the expected dramatic increase of patients affected by dementia and the anticipated costs to the society, efficient prevention strategies – including early imaging markers - are paramount to tackle this major socioeconomic issue. Late-onset Alzheimer’s disease (AD) accounts for more than half of the cases of dementia (Barker et al., 2002) and is characterized by episodic memory impairment in association with executive dysfunctions and visuospatial disturbances (Greene, Hodges, & Baddeley, 1995; Hodges et al., 1999), reflecting amyloid plaques and neurofibrillary tangles deposition in medial temporal and parietal regions (Braak, Thal, Ghebremedhin, & Del Tredici, 2011). While frontotemporal dementia (FTD) is less frequent in the older-age population, its prevalence has been reported as similar to AD in patients aged 45-64 (Ratnavalli, Brayne, Dawson, & Hodges, 2002). FTD encompasses a group of clinical syndromes characterized by behavioural and language changes, due to degeneration in frontal and temporal lobes, with pathologic diagnosis based on abnormal accumulation of three major proteins: microtubule-associated protein tau (MAPT, 40%), TAR DNA-binding protein of 43 kD (TDP-43, 50%) and fused in sarcoma protein (FUS, 10%). FTD includes a behavioural variant (bvFTD) and a primary progressive aphasia (PPA) further divided in a semantic variant (svPPA) and non-fluent type (nfPPA) (Gorno-Tempini et al., 2011; Rascovsky et al., 2011).

Structural MRI imaging studies have identified a distinct pattern of grey matter (GM) atrophy and cortical thinning in AD and FTD (Du et al., 2007; Singh et al., 2006). While medial temporal and parietal lobes are primarily involved in AD, FTD is characterized by cortical thinning that encompasses the frontal and anterior temporal lobes. The FTD subtypes also exhibit anatomical alterations in distinct regional pattern, with cortical thinning more specifically involving the orbitofrontal cortex in bvFTD (Moller et al., 2016), the left anterior temporal cortex in svPPA (Routier et al., 2018) and the inferior frontal gyrus in nfPPA (Agosta et al., 2015). These neuroanatomical patterns also adhere to the clinical syndromes and partly explain the the characteristic clinical and neuropsychological features of each variant.

In addition to cortical thickness, other surface-based morphometry (SBM) indices can characterize cortical folding patterns. One measure is the local gyrification index, defined as the ratio between the inner surface size to the outer surface size of a convex hull. However, this measure suffers from several drawbacks, including between-subject brain size differences normalization and noise in surface reconstruction (Lopes & Betrouni, 2009). Another recently introduced measure of cortical complexity is the fractal dimension, which does not rely on defining an explicit outer hull and thus avoiding the possible confounds encountered when estimating local gyrification index. Based on the idea that the brain structure can be mathematically described as a fractal (Kiselev, Hahn, & Auer, 2003), fractal dimension can be used to measure cortical folding complexity, even at the vertex level (Yotter, Nenadic, Ziegler, Thompson, & Gaser, 2011). Recent studies have shown significant differences of regional fractal dimension in a variety of neuropsychiatric and neurological conditions, including schizophrenia and multiple sclerosis (Esteban et al., 2007; Narr et al., 2004). In AD and mild cognitive impairment (MCI), reduction in fractal dimensino has been observed in the insula, medial temporal lobe and cingulate cortex (Ruiz de Miras et al., 2017). In addition, Sheelakumari *et al*. showed that the whole brain fractal dimension was reduced in behavioural and aphasic variants of FTD compared to controls (Sheelakumari et al., 2018). However, the vertex-wise regional pattern of fractal dimension reductions in FTD has remained largely unknown. Moreover, whereas fractal dimension reduction in AD was related to global cognitive impairment according to the Alzheimer Disease Assessment Scale cognitive (ADAS-cog) scale (King et al., 2010), the correlation between cortical complexity analysis and specific cognitive subdomains has not yet been evaluated.

In this study, we aimed at assessing the regional patterns of cortical thickness and fractal dimension changes in a cross-sectional cohort of patients with AD and FTD. We hypothesized that, in addition to the previously described cortical thinning encompassing the temporoparietal areas in AD and frontotemporal regions in FTD (Du et al., 2007; Moller et al., 2016; Singh et al., 2006; Wang et al., 2009), the two disease groups would exhibit a distinct pattern of fractal dimension reduction. Based on a previous study (Ruiz de Miras et al., 2017), we expected AD subjects to show a reduced fractal dimension in medial temporal regions. As the pattern of altered fractal dimension in AD broadly follows that of cortical thinning, we hypothesized that FTD would present a fractal dimension reduction in disease-specific regions, including the “epicentre” of pathogenesis in the insula (Seeley, 2010) and orbitofrontal cortex.

## 2. METHODS

### 2.1 Participants

The present study is part of the Neuroimaging of Inflammation in MemoRy and Other Disorders (NIMROD) protocol (Bevan-Jones et al., 2017). We included 18 participants with clinically probable AD according to McKhann’s criteria (McKhann et al., 2011), in addition to 14 patients with MCI defined by a Mini Mental State Examination (MMSE) score > 24/30, memory impairment at least 1.5 standard deviation (SD) below that expected for age and education, and *in vivo* evidence of amyloid pathology (positive ^11^C-Pittsburgh compound B (PiB) PET imaging). AD and MCI patients were combined on the basis that these two subgroups represent a clinical continuum of the same pathological spectrum.

We also included 24 patients with FTD (8 bvFTD, 9 svPPA and 7 nfPPA) diagnosed according to published consensus criteria (Gorno-Tempini et al., 2011; Rascovsky et al., 2011). Twenty-eight similarly aged healthy participants were recruited as controls, with MMSE >26/30, absence of regular memory complaints, and no history of major neurological, psychiatric or significant medical illness. Patients were identified from the memory clinic at the Cambridge University Hospitals NHS Trust. Controls were recruited via the Dementias and Neurodegenerative Diseases Research Network (DeNDRON) volunteer register. Informed written consent was obtained in accordance with the Declaration of Helsinki. The study received a favourable opinion from the East of England Ethics Committee (Cambridge Central Research, Ref. 13/EE/0104). Clinical and cognitive assessment included Mini-Mental State Examination (MMSE) and revised Addenbrooke’s Cognitive Examination (ACE-R) (Mioshi, Dawson, Mitchell, Arnold, & Hodges, 2006).

### 2.2 MRI acquisition and preprocessing

Participants underwent MRI imaging acquired on a 3T scanner (Siemens Magnetom Tim Trio) using a magnetization-prepared rapid gradient echo (MPRAGE) T1-weighted sequence with the following parameters: repetition time = 2300 ms, echo time = 2.98 ms, field of view = 240 x 256 mm^2^, 176 slices, flip angle = 9°, isotropic 1mm^3^ voxels.

Surface-based morphometry (SBM) analyses were performed using the Computational Anatomy Toolbox 12 (CAT12, Structural Brain Imaging Group, University of Jena, Germany) in Matlab R2019a version 9.6 (MathWorks Inc., Sherborn, MA, USA). Cortical thickness and central surface of the left and right hemispheres were assessed using a projection-based thickness method (Dahnke, Yotter, & Gaser, 2013). Using a tissue segmentation to estimate the white matter distance, the software projects the local maxima (which is equal to the cortical thickness) to other GM voxels by using a neighbour relationship described by the white matter distance. Projection-based thickness allows the handling of partial volume information, sulcal blurring, and sulcal asymmetries without explicit sulcus reconstruction (Dahnke et al., 2013), which results in a significant reduction of processing time compared to other SBM softwares. Topological correction, spherical mapping and spherical registration are performed in order to obtain vertex-wise cortical thickness. Surface maps were smoothed using a 15mm-full-width-at-half-maximum (FWHM) kernel. Similarly, CAT12 can extract FD values at the global (whole-brain), regional (based on regions of interest of an atlas) and local (vertex) level, based on a spherical harmonic reconstruction method described in a previous study (Yotter et al., 2011). At variance with the box-counting approach, spherical harmonic reconstruction allows to maintain an identical number of vertices for all reconstructed surfaces, which reduces the influence of individual vertex alignment, resampling and interpolation, resulting in more accurate reconstructions (Yotter et al., 2011). After obtaining individual vertex-wise fractal dimension maps, smoothing was performed with a 20mm-FWHM kernel as recommended by CAT12. Labelling of the significant regions of interest determined in group comparisons and correlations was based on the Desikan-Killiany atlas included in the CAT12 SBM toolbox.

### 2.3 Statistical analyses

Demographic data were analyzed with Stata software Version 14.2 (College Station, TX). Assessment of distribution for continuous variables was performed with Shapiro–Wilk test and visualization of histogram plots, followed by ANOVA or Kruskal-Wallis test, and post hoc Tukey/Dunn test, accordingly. Categorical variables were compared with Chi-squared test. Statistical significance was considered when p < 0.05.

Global (whole-brain) cortical thickness and fractal dimension values were calculated for each group, and correlation between cortical thickness and fractal dimension were performed with Pearson correlation. Between-group local (vertex-wise) cortical thickness and fractal dimension comparisons were performed in CAT12 with a 2-sample t-test using age as covariate. Vertex-wise correlations between SBM maps and ACE-R cognitive subscores were performed using multiple regressions with age as covariate. A significant statistical threshold of false discovery rate (FDR)-corrected p < 0.05 was considered for all vertex-wise analyses. We also report exploratory results at p < 0.001 uncorrected.

## 3. RESULTS

### 3.1 Demographics

Demographic and clinical characteristics of patients with AD, FTD and control participants are shown in **Table 1**. A difference in age was observed (p = 0.01, *ANOVA*), with FTD patients being significantly younger than AD patients (p = 0.01, *post hoc Tukey test*), as expected on the basis of the demographic characteristics of these neurodegenerative disorders. Gender distribution and education attainment were similar across groups, whereas, as expected, MMSE and ACE-R scores were significantly lower in the AD and FTD groups compared to Controls (p<0.0001, *Kruskal-Wallis with post hoc Dunn test*). Each of the five ACE-R cognitive subdomains was assessed separately for AD and FTD (including its variants). A significant impairment was considered when the mean group subscore was < 80% of the total maximal subscore. While AD showed an impairment in memory and fluency, FTD group including its variants had a significant degree of impairment in language over and above memory and verbal fluency impairment. Attention/orientation and visuospatial subscores were only mildly reduced in both dementia groups (mean subscores > 80%), and therefore were not used for further analyses of clinical-imaging correlations.

**TABLE 1.**

|  | **AD (n=32)** | **FTD (n=24)** | **Controls (n=28)** | pval |
| --- | --- | --- | --- | --- |
| Age (years) | 72.3 ± 8.2<br>(53-86) | 66.2 ± 9.2<br>(50-84) | 70.3 ± 5.6<br>(59-84) | 0.01 * |
| Male participants | 56.3%<br>(18/32) | 50%<br>(12/24) | 53.6%<br>(15/28) | 0.9 § |
| Education (years) | 13.1 ± 3.0<br>(10-19) | 13.5 ± 2.8<br>(10-18) | 14.6 ± 2.5<br>(10-19) | 0.12 # |
| MMSE | 24.9 ± 3.4<br>(12-30) | 25.2 ± 4.9<br>(14-30) | 29.0 ± 1.0<br>(27-30) | 0.0001 # |
| ACE-R | 75.7 ± 11.0<br>(43-91) | 67.7 ± 17.1<br>(38-93) | 93.7 ± 4.8<br>(79-100) | 0.0001 # |
| Mean CTh | 2.50 ± 0.14\$ | 2.50 ± 0.12 | 2.64 ± 0.10 | 0.0001 * |
| Mean FD | 2.54 ± 0.02\$ | 2.54 ± 0.03 | 2.56 ± 0.02 | 0.0007 * |

Baseline characteristics of included subjects. \*ANOVA, §Chi-squared test, #Kruskall-Wallis

### 3.2 Cortical thickness/FD group comparisons

As shown in **Table 1**, whole-brain cortical thickness and fractal dimension values were significantly lower in AD and FTD relative to controls (p<0.001, *ANOVA* with all *post hoc Tukey test* p<0.001). There was also a significant positive correlation between mean cortical thickness and mean fractal dimension (rho = 0.30, p < 0.006, *Pearson correlation*).

Vertex-wise cortical thickness group comparisons showed that AD subjects had significant cortical thinning in extensive frontal, temporal, parietal and occipital cortices, while FTD patients exhibited cortical thickness reduction mainly in frontal and anterior temporal regions (FDR p < 0.05). Analyses in FTD subgroups revealed that the superior frontal cortex was particularly affected in bvFTD and nfPPA, whereas the left anterior temporal regions were more severely affected in svPPA (all FDR p < 0.05) (**Figure 1**).

**FIGURE 1:**
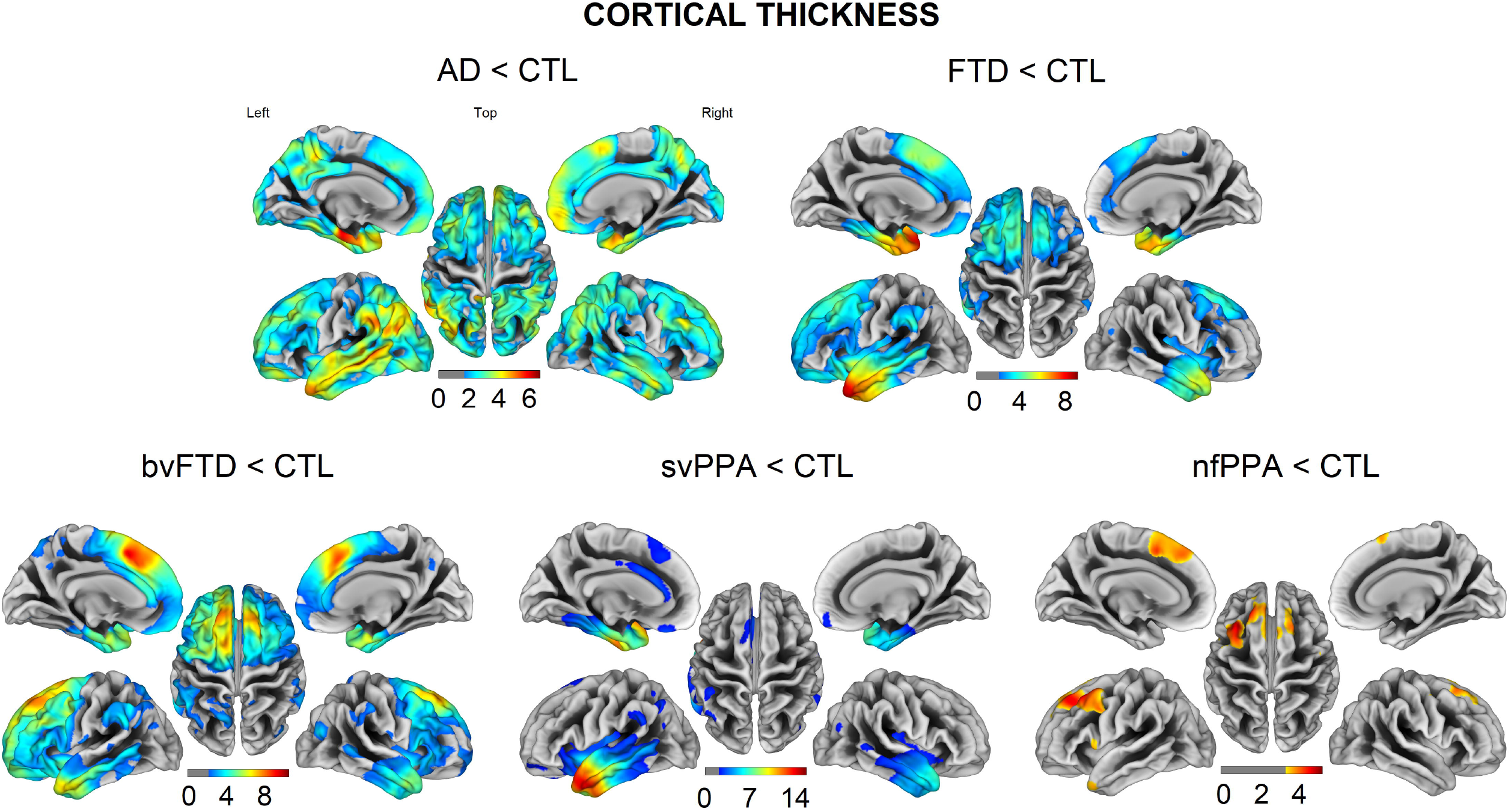
vertex-wise cortical thickness group comparisons between Controls, AD and FTD, including its variants (FDR-corrected p<0.05). Color bars represent T-scores.

We also observed decreased fractal dimension for both AD and FTD relative to controls. More specifically, AD showed reduced fractal dimension in the bilateral insula and supramarginal gyrus, left middle frontal, superior temporal, inferior temporal, and right parahippocampal gyri. FTD patients had reduced fractal dimension in the bilateral insula, middle and inferior frontal, orbitofrontal, as well as left superior parietal, supramarginal and pericalcarine gyri (all FDR p < 0.05). Its variants showed a variable degree of fractal dimension reduction in the insula, orbitofrontal and middle frontal regions (p<0.001) (**Figure 2**). At a more liberal threshold (uncorrected p < 0.001), increased fractal dimension was observed in the anterior frontal regions in AD and lateral temporal areas in FTD.

**FIGURE 2:**
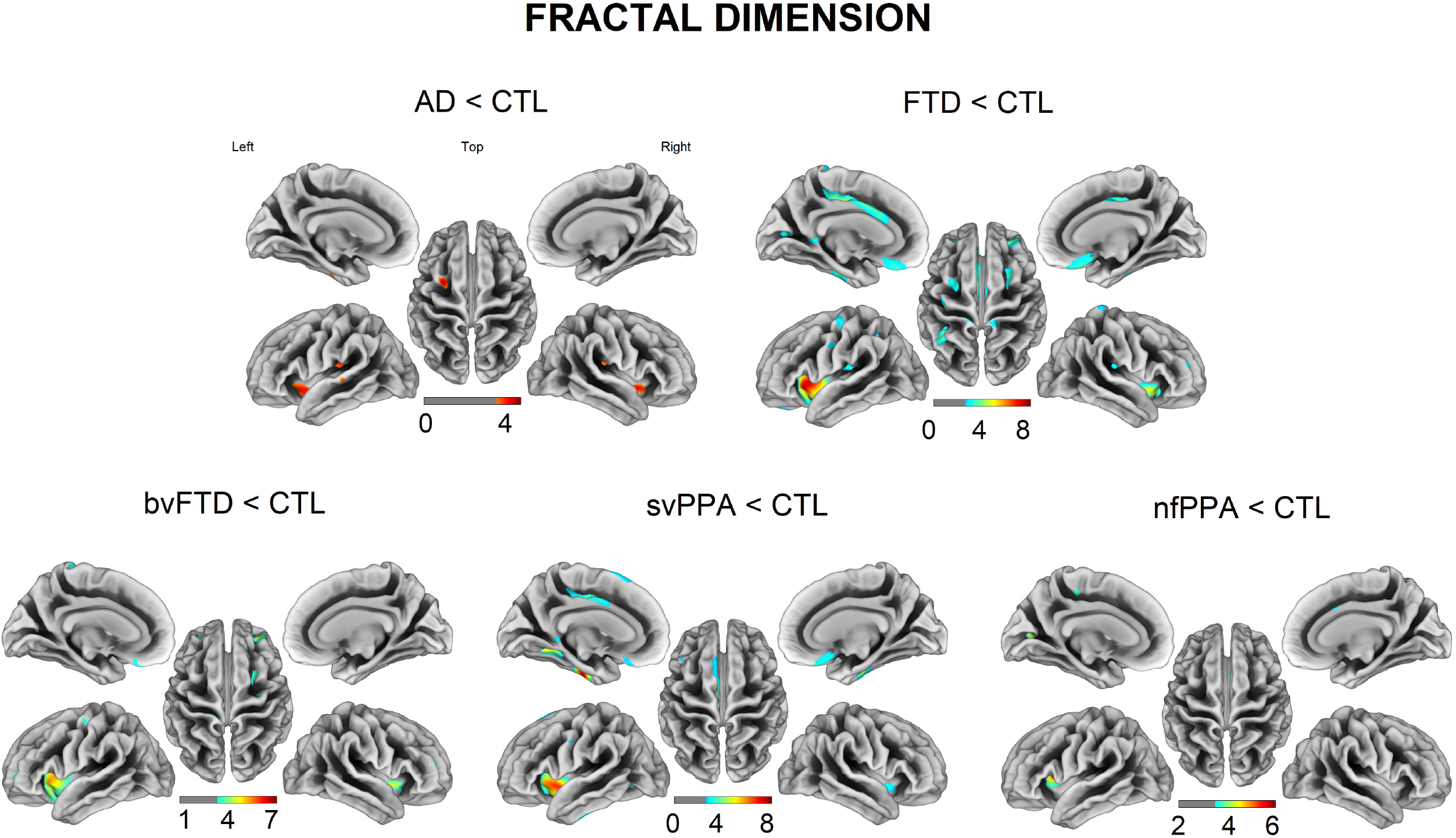
vertex-wise fractal dimension group comparisons between Controls, AD and FTD, including its variants (FDR-corrected p<0.05). Color bars represent T-scores.

Comparisons between AD and FTD revealed decreased cortical thickness for AD in posterior cingulate, parietal and occipital regions, while FTD had cortical thinning in anterior temporal cortices (p<0.001). A significant fractal dimension reduction was observed in the left posterior cingulate and precuneus for AD, while FTD patients had decreased fractal dimension in bilateral orbitofrontal cortex and left insula (p<0.001) (**Figure 3**).

**FIGURE 3:**
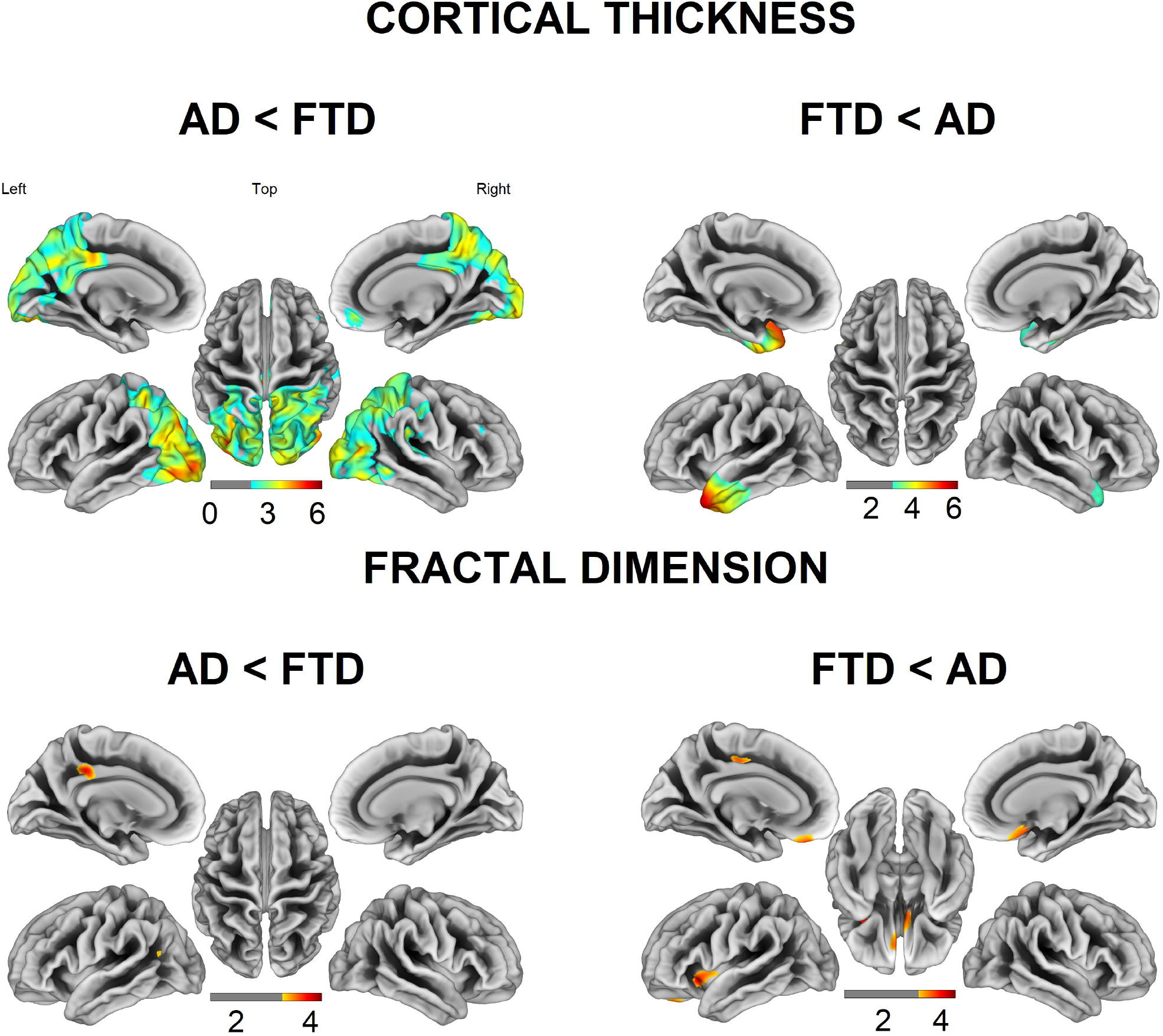
vertex-wise cortical thickness and fractal dimension comparisons between AD and FTD (p<0.001). Color bars represent T-scores.

### 3.3 Vertex-wise correlation of cortical complexity measures and cognition

Memory impairment was correlated with cortical thinning in the left parahippocampal gyrus in AD subjects and in more extensive regions for FTD, including left entorhinal and parahippocampal, left fusiform, posterior cingulate and precuneus, as well as right superior temporal cortex (p < 0.001, uncorrected). Language impairment was associated with a reduced cortical thickness for FTD in left fusiform, superior and inferior temporal, and insula (p < 0.001, uncorrected). No significant correlation was observed in AD for language impairment, as well as for cortical thickness/fluency anatomical correlates in either AD or FTD (**Figure 4**, left-hand side).

**FIGURE 4:**
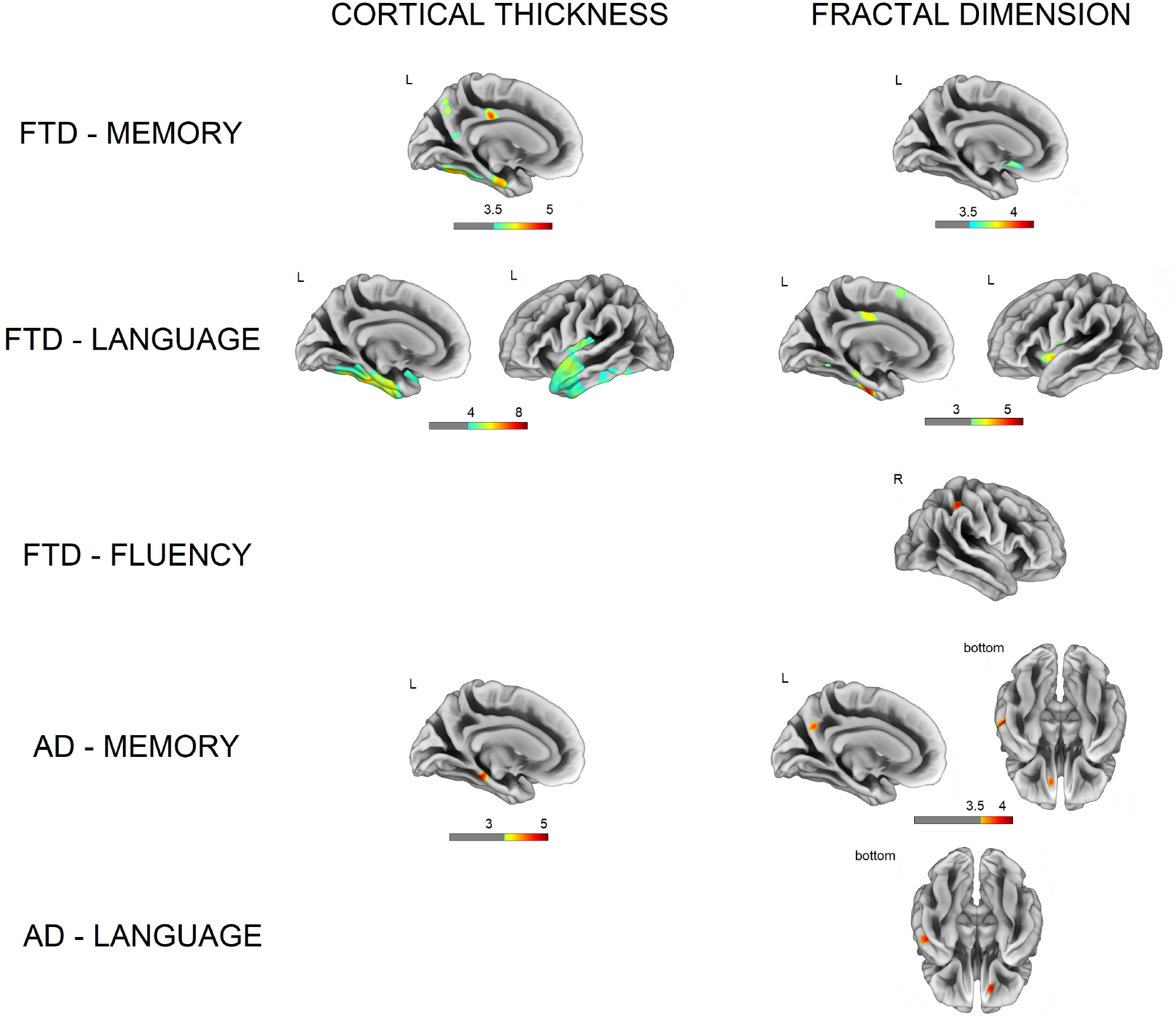
vertex-wise cortical thickness and fractal dimension correlate of memory, fluency and language impairment (according to the ACE-R scale) in AD and FTD (p<0.001). Color bars represent T-scores. L = left, R = right.

Memory impairment was associated with reduced fractal dimension in left middle temporal, inferior temporal, precuneus and medial orbitofrontal for AD and left medial orbitofrontal for FTD, while poor performance in the language subdomain correlated with decreased fractal dimension in the left middle temporal and right lateral orbitofrontal cortices for AD, and in left insula, inferior temporal, posterior cingulate, lingual and parahippocampal gyri regarding FTD. Finally, decreased fluency was associated with reduced fractal dimension in the right superior parietal gyrus in FTD (all p < 0.001, uncorrected), without any significant correlation regarding AD (**Figure 4**, right-hand side).

## 4. DISCUSSION

Neurodegeneration changes cortical complexity. This study demonstrates that the fractal dimension of cortical complexity is a promising imaging tool to assess specific morphological patterns of grey matter damage in degenerative conditions, namely AD and FTD. The fractal dimension in disease-related regions was also related to the severity of cognitive impairment.

We confirmed the typical cortical thinning signature of both AD (temporoparietal regions) and FTD (frontal and anterior temporal regions). In addition, these two conditions have distinct features regarding fractal dimension: whereas both AD and FTD have a variable reduction of fractal dimension in the middle frontal cortex and supramarginal gyrus compared to controls, direct comparisons between groups revealed that precuneus and posterior cingulate regions were particularly vulnerable in AD, while orbitofrontal gyrus and insula showed a more pronounced fractal dimension reduction for FTD subjects. Recently, Ruiz de Miras *et al*. showed that white matter fractal dimension (but not pial surface fractal dimension) was reduced for AD subjects in medial temporal lobe, insula and posterior cingulate (Ruiz de Miras et al., 2017). Using a similar spherical harmonic reconstruction proposed by Yotter (Yotter et al., 2011) natively embedded in the CAT12 toolbox, we were able to observe similar regional changes at the cortical level.

Our identification of vertex-wise changes in fractal dimensionality for FTD differs from Sheelakumari *et al*. who found a decrease in fractal dimension at the whole-brain and hemispheric level: a decrease in the general fractal structure was observed for bvFTD, while PPA subjects had more prominent impairment in the left hemisphere. We found that FTD had reduced fractal dimension in the insula, middle and inferior frontal, orbitofrontal and posterior regions compared to controls. In addition, we repeated these analyses for the different FTD variants and observed that inferior frontal and insula were impaired in all groups. bvFTD and svPPA variants showed fractal dimension reductions in orbitofrontal areas, while only svPPA had a decreased fractal dimension in (mostly left) parahippocampal cortex. These findings support the insula as being a major hub in speech production and socio-emotional functioning (Mandelli et al., 2016; Seeley, 2010).

When comparing the SBM maps of AD/FTD patients with controls, we observed that cortical thickness represented a more sensitive measure to detect cortical changes in dementia. In fact, large clusters of cortical areas showing a decreased cortical thickness in AD and FTD were observed (**Figure 1**), while fractal dimension changes were mostly localised in limbic and cingulate areas (**Figure 2**).

One explanation could be that fractal dimension can decrease or increase in degenerative conditions according to how the structural impairment involves the pial surface. In fact, a change in the pial surface decreasing the folding area would more likely decrease complexity. Conversely, if the change involves an increase in sulcal depth, the complexity (and thus fractal dimension) would increase. While alterations in both directions were observed by King *et al*. (King et al., 2010) using whole-brain fractal dimension analyses, we failed to observe a significant increase in the two dementia groups with a FDR-corrected threshold.

While our study suggests that fractal dimension is a less sensitive structural marker in comparison to cortical thickness, we observed that the cortical complexity changes in fractal dimensionality correlated well with cognitive subdomains impaired in AD and FTD. In fact, while memory impairment was mainly associated with cortical thinning in parahippocampal regions, we observed additional correlational features for fractal dimension, e.g. a reduction in orbitofrontal cortex in both AD and FTD (**Figures 3-4**). These findings add credence to the hypothesis that orbitofrontal area, whose rostral region is primarily linked to medial temporal limbic structures, plays a major role in memory encoding (Duarte, Henson, Knight, Emery, & Graham, 2010; Frey & Petrides, 2000).

The study has several limitations. First, its cross-sectional design impeded further analyses regarding how cortical complexity evolves longitudinally. In addition, we enrolled FTD subjects at the dementia stage, while the AD group included patients with clinically probable AD as well as MCI with biomarker evidence of amyloid pathology. Further studies should assess fractal dimension changes in earlier cases, e.g. presymptomatic mutation carriers, especially in FTD. This would test whether MRI-based fractal dimension is a sensitive measure to detect early cortical alterations. Finally, our FTD subtype analysis had small group sizes, limiting power, although the significant results were convergent in the insula across the different subtypes.

This study has identified the impact of AD and FTD pathologies on cortical patterns of fractal dimensionality. Further work will determine when these changes emerge and how quickly they progress, in relation to other biomarkers, and in relation to the cellular and molecular features of neurodegenerative diseases.

## Data Availability

The data that support the findings of this study are available from the corresponding author upon reasonable request.

## ACKNOWLEDGEMENTS

Thanks to our volunteers for participating in this study and to the radiographers at the Wolfson Brain Imaging Centre, University of Cambridge, UK, for their invaluable support in data acquisition. We thank the NIHR Dementias and Neurodegenerative Diseases Research Network for help with subject recruitment.

## Compliance with ethical standards

### Ethical approval

The present study was performed in agreement with the Declaration of Helsinki and its further amendments. Approval was obtained from Ethics Committee from East of England (Cambridge Central Research, Ref. 13/EE/0104).

### Informed consent

Informed consent has been obtained from all participants in the present study.

